# Muscle strength is associated with COVID-19 hospitalization in adults 50 years of age or older

**DOI:** 10.1101/2021.02.02.21250909

**Authors:** Boris Cheval, Stefan Sieber, Silvio Maltagliati, Grégoire P. Millet, Tomáš Formánek, Aïna Chalabaev, Stéphane Cullati, Matthieu P. Boisgontier

## Abstract

**Background:** Weak muscle strength has been associated with a wide range of adverse health outcomes. Yet, whether individuals with weaker muscle strength are more at risk for hospitalization due to severe COVID-19 is still unclear. The objective of this study was to investigate the independent association between muscle strength and COVID-19 hospitalization.

**Methods:** Data from adults 50 years of age or older were analyzed using logistic models adjusted for several chronic conditions, body-mass index, age, and sex. Hand-grip strength was repeatedly measured between 2004 and 2017 using a handheld dynamometer. COVID-19 hospitalization during the lockdown was self reported in summer 2020 and was used as an indicator of COVID-19 severity.

**Results:** The study was based on the Survey of Health, Ageing and Retirement in Europe (SHARE) and included 3600 older adults (68.8 ± 8.8 years, 2044 females), amongst whom 316 were tested positive for the severe acute respiratory syndrome coronavirus 2 (8.8%) and 83 (2.3 %) were hospitalized due to COVID-19. Results showed that higher grip strength was associated with a lower risk of COVID-19 hospitalization (adjusted odds ratio [OR] per increase of 1 standard deviation in grip strength = .64, 95% confidence interval [95% CI] = .45–.87, *p* = .015). Results also showed that age (OR for a 10-year period = 1.70, 95% CI = 1.32–2.20, *p* < .001) and obesity (OR = 2.01, 95% CI = 1.00–3.69, *p* = .025) were associated with higher risk of COVID-19 hospitalization. Sensitivity analyses using different measurements of grip strength as well as robustness analyses based on rare-events logistic regression and a different sample of participants (i.e., COVID-19 patients) were consistent with the main results.

**Conclusion:** Muscle strength is an independent risk factor for COVID-19 severity in adults 50 years of age or older.

## Introduction

As of February 2021, more than 100 million people were diagnosed with the coronavirus disease 2019 (COVID-19) and over 2 million died due to this infection (1). The majority of infected people are asymptomatic (2, 3) or have mild symptoms such as fever, cough, dyspnea, fatigue, or anosmia/dysgeusia (4, 5). However, severe COVID-19 symptoms can also be life-threatening and require hospitalization (6). Thus, identifying risk factors for severe COVID-19 is important to inform clinical decisions and public-health strategies.

Several risk factors have already been identified, including older age, male sex, as well as underlying health conditions such as obesity, cardiovascular disease, respiratory disease, kidney disease, diabetes, and cancer (7-9). In addition to these established risk factors for severe COVID-19, the latest studies suggest that physical fitness should also be considered (10-12). For example, maximum exercise capacity was associated with the risk of COVID-19 hospitalization (11); slower walkers showed higher risk of severe COVID-19 than brisk walkers (12), and the overall level of fitness was associated with survival in COVID-19 hospitalized patients (13). Whether muscle strength, another dimension of physical fitness, is a risk factor for severe COVID-19 remains unclear.

Muscle strength is an indicator of muscle function, which is essential to health (14, 15) and has shown to be a robust predictor of multiple diseases and all-cause mortality (14, 16-18). Therefore, muscle strength should be considered as a potential risk factor for severe COVID-19 (19). The objective of this study was to investigate the association between muscle strength and COVID-19 severity. We hypothesized that maximum muscle strength would be inversely associated with COVID-19 hospitalization, independently of established risk factors.

## Methods

### Study overview

Data from the Survey of Health, Ageing and Retirement in Europe (SHARE) were collected every two years between 2004 and 2017 (7 waves of data collection) on adults 50 years of age or older living in 27 European countries (n = 139556). From June to September 2020, SHARE participants (n = 52310) responded to additional questions in the SHARE COVID-19 questionnaire (20). Questions included whether they had been tested positive for the severe acute respiratory syndrome coronavirus 2 (SARS-CoV-2) and whether this infection had resulted in hospitalization. To be included in the study, participants should be aged 50 years or older, have completed at least one regular (not COVID-19) SHARE questionnaire between 2004 and 2017. They should also have indicated whether they were infected by the SARS-CoV-2 and/or whether they were hospitalized due to COVID-19 in the SHARE COVID-19 questionnaire (Figure 1). SHARE was approved by the Ethics Committee of the University of Mannheim (waves 1-4) and the Ethics Council of the Max Plank Society (waves 4-7).

**Figure 1.**
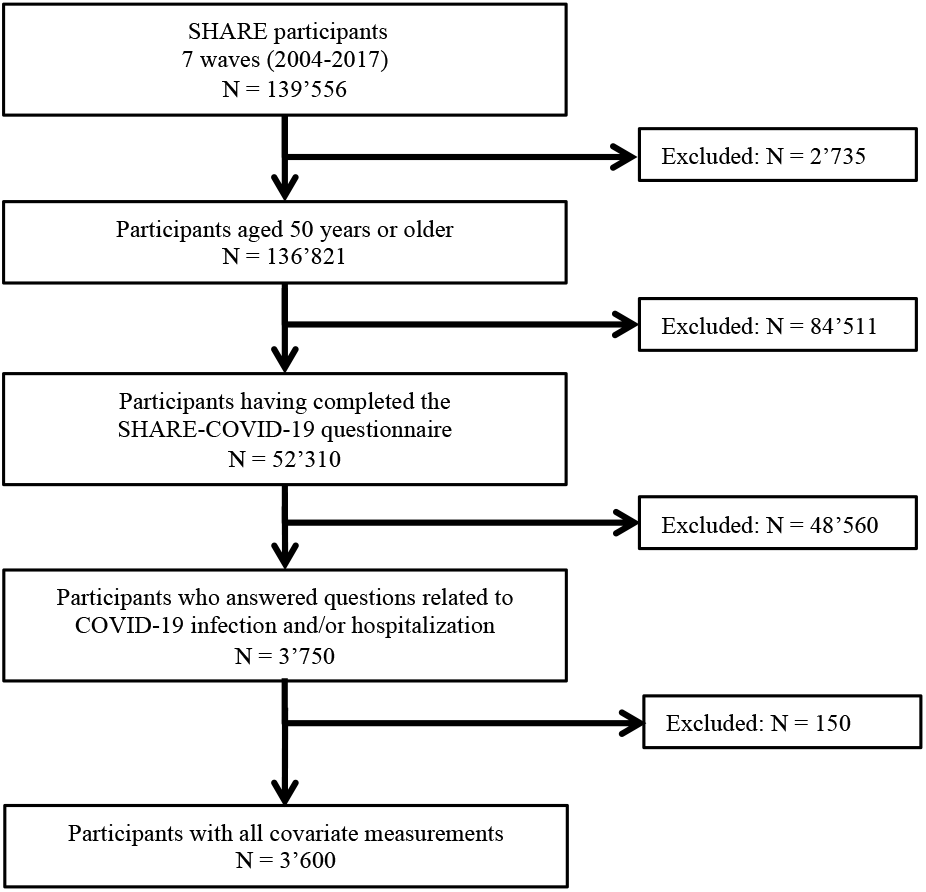
Flow Chart

### Measures

#### Outcome: COVID-19 hospitalization

Hospitalization due to COVID-19 was derived from the question: “*Have you, or anyone close to you, been hospitalized due to an infection from the coronavirus?*”. If participants answered “*yes*”, the interviewer asked who was hospitalized. Participants who indicated they were hospitalized were included in the analyses as COVID-19 hospitalized. If the participant indicated that their “*spouse or partner*” was hospitalized, the spouse or partner was included in the analyses as COVID-19 hospitalized.

#### Independent variable: Muscle strength

Hand-grip strength (kg) was used as an indicator of muscle strength and was measured twice with each hand (alternating between hands) using a handheld dynamometer (Smedley, S Dynamometer, TTM, Tokyo, 100 kg). Participants were instructed to stand (preferably) or sit with the elbow flexed at a 90° angle, the wrist in a neutral position, and the upper arm in a vertical position against the trunk. Interviewers delivered standardized instructions to ensure that the grip was performed with maximum effort. The maximum value collected in either hand was used as an indicator of muscle strength in the main analyses (14, 21). Grip strength was assessed at each data-collection wave, but only the most recent measurement of maximum grip strength was included in the main analyses.

#### Covariates and established risk factors

The following covariates were included in the analysis: *Age* (in 2020, when responding to the SHARE COVID-19 questionnaire), *sex* (male, female), *height* (cm), *body-mass index* (normal: <25, overweight: ≥25 and <30, obese: ≥30 kg/m^2^), *cardiovascular disease* (heart attack, including myocardial infarction or coronary thrombosis, or any other cardiovascular problem including congestive heart failure, high blood cholesterol, high blood pressure or hypertension, stroke or cerebral vascular disease), *respiratory disease* (including chronic bronchitis or emphysema, and asthma), *diabetes, cancer, chronic kidney disease*, and *rheumatoid arthritis*. All these covariates are established risk factors for severe COVID-19 (7-9) and were measured using questionnaires. When a participant had multiple measures across the different waves of data collection, the most recent measurement was included in the analyses. As recommended (17, 22, 23), self-reported height (cm) was included in the analyses to ensure that the association observed between muscle strength and COVID-19 hospitalization was not due to a difference in height.

### Data analyses

#### Main analyses

Three logistic regression models were fitted. Model 0 tested the association between muscle strength and COVID-19 hospitalization, adjusting for height. Model 1 tested the association between the established risk factors (i.e., age, sex, height, body-mass index, cardiovascular disease, respiratory disease, diabetes, cancer, chronic kidney disease, and rheumatoid arthritis) and COVID-19 hospitalization. Model 2 tested the association between grip strength and COVID-19 hospitalization, while adjusting for the established risk factors. Age was centered on mean age (i.e., 68.8 years) and divided by 10, so that the coefficient reflected the effects of increased odds of COVID-19 hospitalization over a 10-year period (24). Grip strength was standardized so that the coefficient reflected the effects associated with an increase of 1 standard deviation (SD). The multiple logistic regression models account for the variance shared between muscle strength, age, and health-related conditions, thereby assessing whether muscle strength was independently associated with COVID-19 hospitalization. In other words, the models examined the association between muscle strength and COVID-19 hospitalization over and above the influence of the established risk factors for severe COVID-19 (i.e., age, sex, height, body-mass index, cardiovascular disease, respiratory disease, diabetes, cancer, chronic kidney disease, and rheumatoid arthritis). Statistical analyses were conducted in R using the glm package. Statistical assumptions associated with general logistic models were met (i.e., normality of the residuals, multicollinearity, and undue influence). To illustrate the association between grip strength and COVID-19 hospitalization from the estimates obtained in Model 2, we computed the odds ratio of hospitalization in individuals with weaker grip strength and stronger grip strength by centering grip strength on mean – 1 (SD) and mean + 1SD, respectively.

#### Sensitivity analyses

Three sensitivity analyses were conducted. In the first sensitivity analysis, average grip strength over the study duration (i.e., from wave 1 to 7) replaced the most recent measurement to test the association with a more stable level of muscle strength. The second sensitivity analysis included grip strength assessed only in the wave preceding the SHARE COVID-19 questionnaire (i.e., wave 7) (N = 2,884) to shorten the time between the measure of grip strength and the COVID-19 hospitalization event. The third sensitivity analysis adjusted for the country of residence (Belgium, Bulgaria, Croatia, Cyprus, Czech Republic, Denmark, Estonia, Finland, France, Germany, Greece, Hungary, Israel, Italy, Latvia, Lithuania, Luxembourg, Malta, Netherlands, Poland, Portugal, Romania, Slovakia, Slovenia, Spain, Sweden, Switzerland).

#### Robustness analyses

Two robustness analyses were conducted. In the first robustness analysis, the dataset was analyzed using a rare-events logistic regression (25), which corrects for the bias associated with rare events. To account for the estimated fraction of patients hospitalized due to COVID-19 in the European population from June to September 2020, we used a tau parameter of 84/100000 based on COVID-19 hospitalization data that were available from May 2020 (Table S1). These data likely underrepresent the true number of COVID-19 hospitalizations, as hospitalizations that occurred earlier than May 2020 were not included, thereby making the analysis more conservative. In addition, we corrected for our case-control sampling design using the weighting method (R Zelig package) (26). The second robustness analysis included only patients who tested positive for COVID-19 (N = 289) to examine whether muscle strength is a risk factor in the population of COVID-19 patients. This subsample was generated based on the question: “*Have you, or anyone close to you, been tested for the coronavirus and the result was positive, meaning that you or the person had the COVID disease?*”.

## Results

The study sample included 3,600 individuals (68.8 ± 8.8 years, 2044 females), of whom 316 were tested positive for the SARS-CoV-2 (8.8%) and 83 (2.3%) were hospitalized due to COVID-19 (Figure 1). Table 1 summarizes the characteristics of the participants stratified by COVID-19 hospitalization status. COVID-19 hospitalization (vs. no hospitalization) was associated with older age (*p* < .001), higher body-mass index (*p* = .062), cardiovascular disease (*p* = .064), chronic kidney disease (*p* = .052), and weaker muscle strength (*p* = .027). The number of participants who completed their last measure of grip strength in wave 1, 2, 3, 4, 6, and 7 was 10, 7, 3, 39, 188, 413, and 2940, respectively.

**Table 1.**
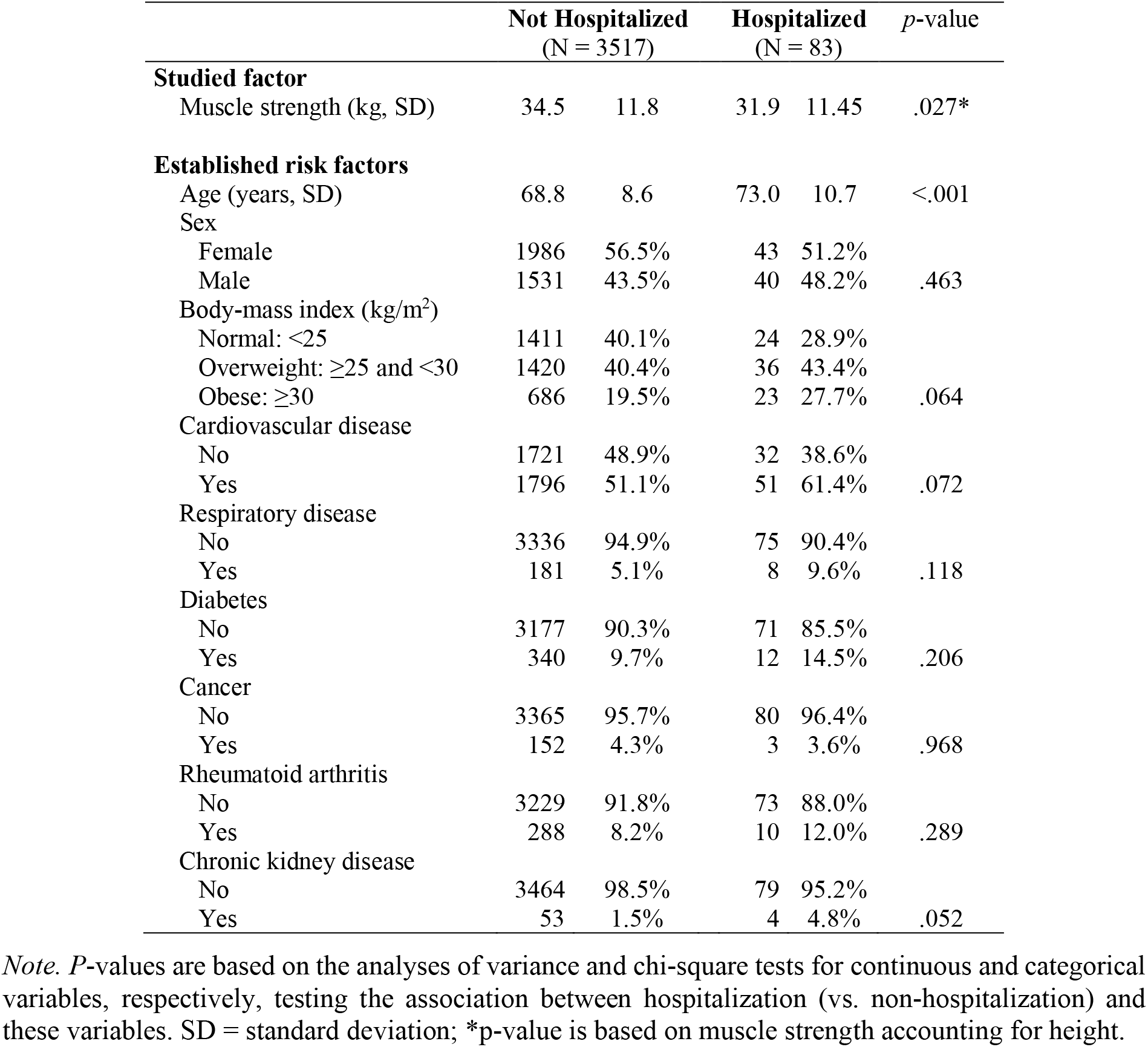
Sample characteristics by hospitalization status

### Univariate model

Model 0 showed that the most recent measure of maximum grip strength was associated with COVID-19 hospitalization (adjusted odds ratio [OR] = .60, 95% confidence interval [95% CI] = .45–.81, *p* < .001, per increase of 1 SD in grip strength) (Table 2).

**Table 2.** Results of the logistic models testing the association of established risk factors and grip strength with COVID-19 hospitalization

|  | Model 0 |  | Model 1 |  | Model 2 |  |
| --- | --- | --- | --- | --- | --- | --- |
| Variables | OR (95% CI) | *p* | OR (95% CI) | *p* | OR (95% CI) | *p* |
| Intercept | .02 (.02; .03) | <.001 | .015 (.01; .02) | <.001 | .012 (.01; .02) | <.001 |
| Hand-grip strength | .60 (.45; .81) | <.001 |  |  | .64 (.45; .92) | .015 |
| Height | 1.54 (1.15; 2.04) | .003 | 1.31 (.96; 1.77) | .089 | 1.06 (1.06; 2.00) | .020 |
| Age |  |  | 1.70 (1.32; 2.20) | <.001 | 1.50 (1.14; 1.97) | .003 |
| Sex (ref. Female) |  |  |  |  |  |  |
| Male |  |  | .79 (.43; 1.43) | .427 | 1.26 (.62; 2.51) | .518 |
| Body-mass index (ref. Normal) |  |  |  |  |  |  |
| Overweight |  |  | 1.44 (.85; 2.49) | .179 | 1.53 (.90; 2.64) | .121 |
| Obese |  |  | 2.01 (1.09; 3.69) | .025 | 2.11 (1.14; 3.88) | .016 |
| Cardiovascular disease (ref. No) |  |  |  |  |  |  |
| Yes |  |  | 1.06 (.66; 1.73) | .811 | 1.03 (.64; 1.67) | .912 |
| Respiratory disease (ref. No) |  |  |  |  |  |  |
| Yes |  |  | 1.52 (.66; 3.08) | .278 | 1.50 (.65; 3.05) | .295 |
| Diabetes (ref. No) |  |  |  |  |  |  |
| Yes |  |  | 1.19 (.59; 2.19) | .601 | 1.09 (.54; 2.01) | .806 |
| Cancer (ref. No) |  |  |  |  |  |  |
| Yes |  |  | .67 (.16; 1.85) | .504 | .63 (.15; 1.74) | .438 |
| Rheumatoid arthritis (ref. No) |  |  |  |  |  |  |
| Yes |  |  | 1.17 (.55; 2.24) | .669 | 1.07 (.50; 2.07) | .850 |
| Chronic kidney disease (ref. No) |  |  |  |  |  |  |
| Yes |  |  | 2.44 (.71; 6.41) | .104 | 2.29 (.66; 6.05) | .134 |

*Note.* Age was centered on mean age (i.e., 68.8 years) and divided by 10 so that the coefficient reflected the effects of an increased odds of COVID-19 hospitalization over a 10-year period. Hand-grip strength was standardized so that the coefficient reflected the effects associated with an increase of 1 standard deviation. OR = Odds Ratio; 95% CI = 95% Confidence Interval.

### Established risk factors and COVID-19 hospitalization

Model 1 showed that older individuals were at higher risk of COVID-19 hospitalization compared to younger individuals (OR = 1.70, 95% CI = 1.32–2.20, *p* < .001). The odds ratio was also higher in obese individuals than in individuals with a normal body-mass index (OR = 2.01, 95% CI = 1.0–3.69, *p* = .025). The other associations were not statistically significant (*p*s > .089) (Table 2).

### Grip strength and COVID-19 hospitalization

Model 2 showed that the most recent measure of maximum grip strength (34.43 ± 11.79 kg; mean ± SD) was associated with the risk of COVID-19 hospitalization (OR = .64, 95% CI = .45–.92, *p* = .015 per increase of 1 SD in grip strength) (Table 2). The odds ratio was more than twice higher in individuals with weaker grip strength (mean − 1SD = 22.64 kg, OR = .019, 95% CI = .01–.03, *p* < .001), compared to individuals with stronger grip strength (mean + 1SD = 46.22 kg, OR = .008, 95% CI = .003–.016, *p* < .001) (Figure 2). When grip strength was included in the model, the association of age (OR = 1.50, 95% CI = 1.14–1.97, *p* = .003) and body-mass index (OR = 2.11, 95% CI = 1.14–3.88, *p* = .016) with COVID-19 hospitalization remained significant (Table 2).

**Figure 2.**
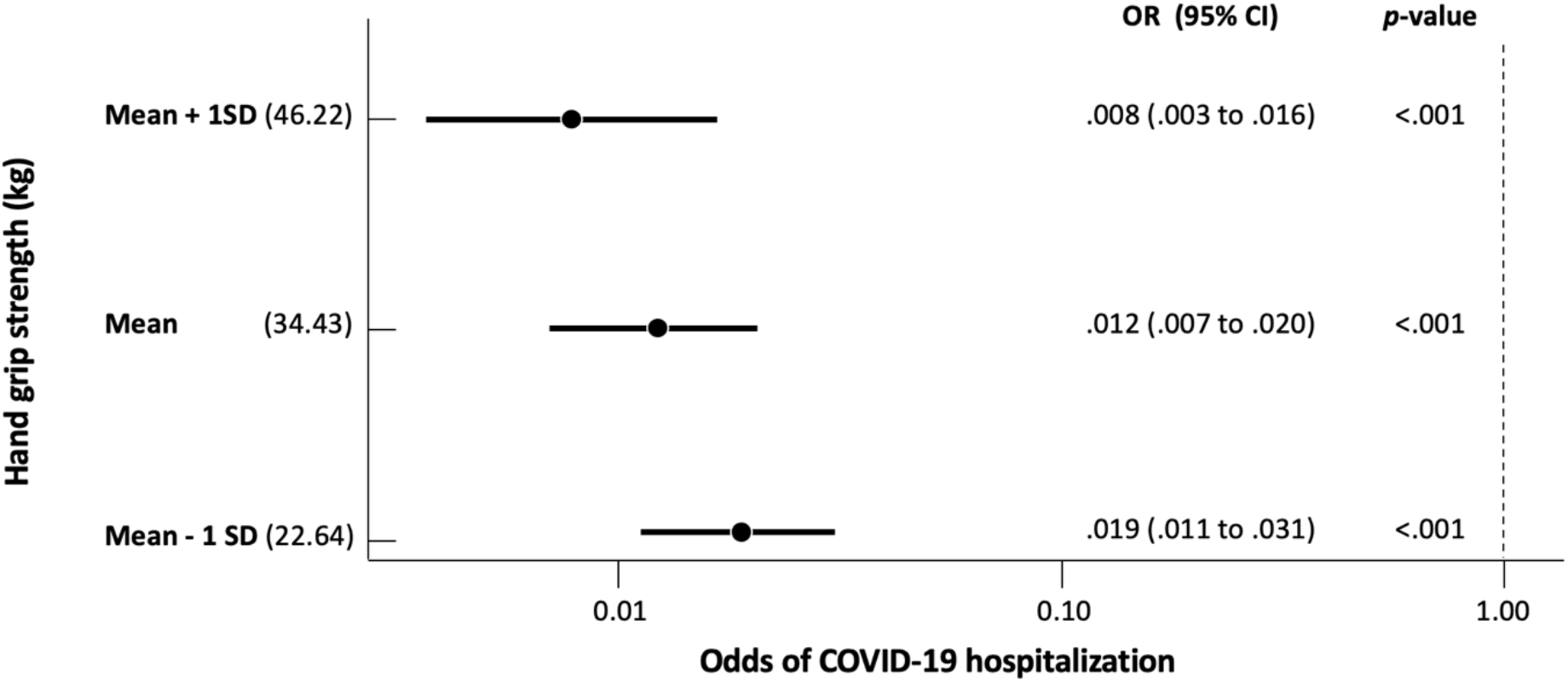
Association between hand-grip strength and the odds of being hospitalized due to COVID-19 *Note*. Odds ratios (OR) were adjusted for age, sex, height, body-mass index, cardiovascular disease, respiratory disease, chronic kidney disease, rheumatoid arthritis, diabetes, and cancer (see Model 2). 95% CI = 95% confidence interval.

### Sensitivity and robustness

The sensitivity (Table S2, S3, and S4) and robustness analyses (Table S5 and S6) yielded similar results to those of the main analyses. Specifically, results of the sensitivity analyses showed that muscle strength averaged across the 7 waves (OR = .62, 95% CI = .42–.92, *p* = .017), muscle strength assessed only in wave 7 (OR = .65, 95% CI = .44–.96, *p* = .029), and the most recent measure of maximum grip strength in the model adjusting for the country of residence (OR = .67, 95% CI = .46–.98, *p* = .0367) were associated with the risk of COVID-19 hospitalization. Results of the robustness analysis based on the rare-events logistic regression showed that the most recent measure of maximum grip strength was associated with the risk of COVID-19 hospitalization (OR = .63, 95% CI = .43–.92, *p* = .016). Finally, results of the robustness analysis that included only patients who tested positive for COVID-19 showed that the most recent measure of maximum grip strength was associated with the risk of COVID-19 hospitalization (OR = .56, 95% CI = .33–.94, *p* = .031).

## Discussion

Recent findings suggest that physical fitness should be considered as a risk factor for severe COVID-19 (10-12). Here, muscle strength was assessed using hand-grip strength and severe COVID-19 was derived from self-reported COVID-19 hospitalization. Results showed that weaker muscle strength was associated with a higher risk of severe COVID-19, which lends direct empirical support to this hypothesized relationship. Moreover, this association was observed after adjusting for established risk factors for severe COVID-19. The latter result suggests that muscle strength was associated with COVID-19 hospitalization, independent of older age, male sex, and health-related conditions including obesity, cardiovascular disease, respiratory disease, diabetes, cancer, chronic kidney disease, and rheumatoid arthritis.

The association between muscle strength and COVID-19 severity can be explained by the essential role of muscle in health and disease (15). In particular, skeletal muscle weakness has been shown to affect the motor and respiratory function, and has been linked to poor immune response and metabolic stress when facing acute infection (19, 27-29). Therefore, adults with weaker muscle strength may be more vulnerable to SARS-CoV-2 infection and at higher risk of developing severe forms of COVID-19. This hypothesis is indirectly supported by previous results. For example, some conditions associated with muscle weakness such as older age, chronic disease, and cancer have been identified as risk factors for COVID-19 severity (30-32). Likewise, patients with sarcopenia have been associated with an impaired respiratory function (33), which is the function affected by SARS-CoV-2. Finally, our findings are consistent with recent studies showing that other dimensions of physical fitness are associated with an increased risk of COVID-19 hospitalization (10-13). Of note, in older patients, the SARS-CoV-2 infection has been shown to increase the risk of muscle loss and acute sarcopenia (19, 34, 35). Therefore, a vicious cycle between muscle weakness and COVID-19 could occur, with muscle weakness increasing COVID-19 severity, which in turn could trigger muscle-mass loss and muscle-strength decline (19), thereby negatively affecting the patients’ condition.

Among the strengths of the present study are the large sample size, the longitudinal design, and a measure of hand-grip strength based on a well-established procedure. Moreover, the results were consistent across multiple independent variables, different statistical approaches, and two different population samples (i.e., general population and patients tested positive for COVID-19). However, potential limitations should be noted. First, the established risk factors were assessed with questionnaires, which may have reduced measurement validity. Second, the latest assessment of these factors was in 2017, two years before participants’ potential COVID-19 infection. Therefore, participants may have contracted a disease sometime between the assessment of these heath conditions and the COVID-19 pandemic, which may have resulted in a misclassification bias. Third, COVID-19 severity was inferred by COVID-19 hospitalization. However, this measure lacks sensitivity. For example, the questionnaire did not assess the duration of hospitalization or whether the patients were transferred to an intensive-care unit, which would have allowed a more nuanced assessment of COVID-19 severity. Similarly, our sample did not include participants who were hospitalized during data collection (except when the information could be extracted from the spouse or partner) and participants who died due to COVID-19. These limitations may explain the absence of statistical evidence supporting the effect of established risk factors for COVID-19 hospitalization (36, 37).

## Conclusion

This study shows that muscle strength is independently associated with the risk of severe COVID-19 in adults 50 years of age or older. These findings further highlight muscle strength as an important factor to monitor in COVID-19 patients (19, 34, 38), thereby suggesting that hand-grip strength could improve the accuracy of composite scores used to predict COVID-19 severity (19, 39, 40).

## Data Availability

This SHARE dataset is available at http://www.share-project.org/data-access.html.

## Statement of Conflict of Interest and Adherence to Ethical Standards

All authors declare that they have no conflict of interests.

## Author Contributions

All the authors designed the study. S.S. cleaned the data. B.C. analyzed the data. B.C., M.P.B. drafted the manuscript. All authors critically appraised the manuscript, worked on its content, and approved its submitted version.

## Ethical approval

This study was part of the SHARE study, approved by the relevant research ethics committees in the participating countries

## Inform Consent

All participants provided written informed consent.

## Funding

B.C. is supported by an Ambizione grant (PZ00P1_180040) from the Swiss National Science Foundation (SNSF).

## Data sharing

This SHARE dataset is available at http://www.share-project.org/data-access.html.

## Acknowledgements

*This paper uses data from SHARE Waves 1, 2, 3 (SHARELIFE), 4, 5,6, 7 and 8 (DOIs: <u>10..6103/SHARE.w1.600, 10..6103/SHARE.w2.600, 10..6103/SHARE.w3.600, 10..6103/SHARE.w4.600, 10..6103/SHARE.w5.600, 10..6103/SHARE.w6.600, 10.6103/SHARE.w7.711, 10.6103/SHARE.w8cabeta.001</u>)*. The SHARE data collection was primarily funded by the European Commission through FP5 (QLK6-CT-2001-00360), FP6 (SHARE-I3: RII-CT-2006-062193, COMPARE: CIT5-CT-2005-028857, SHARELIFE: CIT4-CT-2006-028812) and FP7 (SHARE-PREP: no.211909, SHARE-LEAP: no.227822, SHARE M4: no.261982). Additional funding from the German Ministry of Education and Research, the Max Planck Society for the Advancement of Science, the U.S. National Institute on Aging (U01_AG09740-13S2, P01_AG005842, P01_AG08291, P30_AG12815, R21_AG025169, Y1-AG-4553-01, IAG_BSR06-11, OGHA_04-064, HHSN271201300071C) and from various national funding sources is gratefully acknowledged (see www.share-project.org).

## Notes

### Competing Interest Statement

The authors have declared no competing interest.

### Author Declarations

SHARE was approved by the Ethics Committee of the University of Mannheim(waves 1-4) and the Ethics Council of the Max Plank Society(waves 4-7).

